## Supplementary Materials for "The female protective effect against autism spectrum disorder"

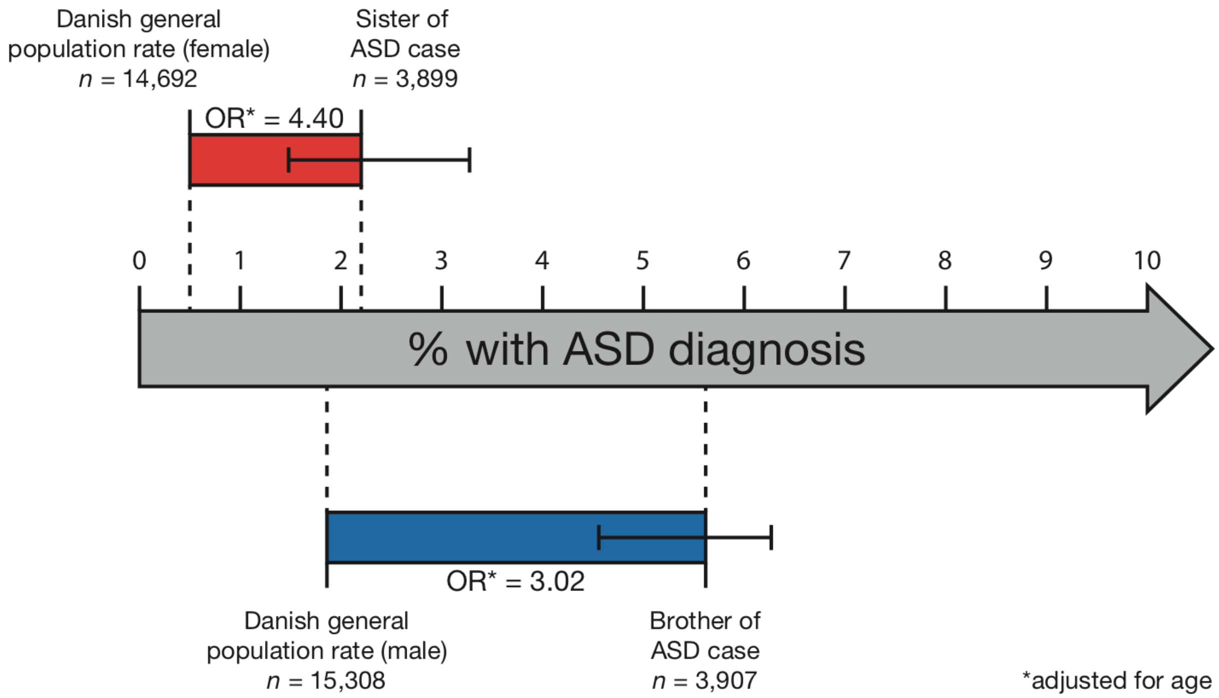


**Supplementary Figure 1. Increased risk for ASD in sisters and brothers of ASD cases compared to Danish population controls.** ORs are the exponentiated betas from logistic regression. Error bars are 95% confidence intervals. The start positions of the coloured bars represent the prevalence of ASD in the Danish general population, by sex. The end positions of the coloured bars represent the projected risk of ASD in siblings, by sex. The end positions are calculated by multiplying the baseline prevalence by the OR.

We hypothesized that given a FPE, brothers of ASD cases would have increased risk for ASD compared to sisters of ASD cases. Sisters of ASD cases have significantly increased risk for ASD (OR = 4.40, 95% CI: 2.96-6.55), calculated as fold-change over age and sex matched controls, compared to brothers of ASD cases (OR = 3.02, 95% CI = 2.45-3.73, $P = 1.75 \times 10^{-9}$, Wald test; see Online Methods). However, the baseline prevalence of ASD amongst females in the Danish general population is lower than the baseline prevalence of ASD amongst males in the Danish general population. Therefore, sisters of ASD cases' overall risk for ASD remains lower than for brothers of ASD cases. The prevalence of ASD in the female Danish general population is 0.5%. A 4.4 fold increase in risk for ASD with a baseline risk of 0.5% would result in a 2.2% chance of having ASD. The prevalence of ASD in the male Danish general population is 1.86%. A 3.02 fold increase in risk for ASD with a baseline risk of 1.86% would result in a 5.62% chance of having ASD.

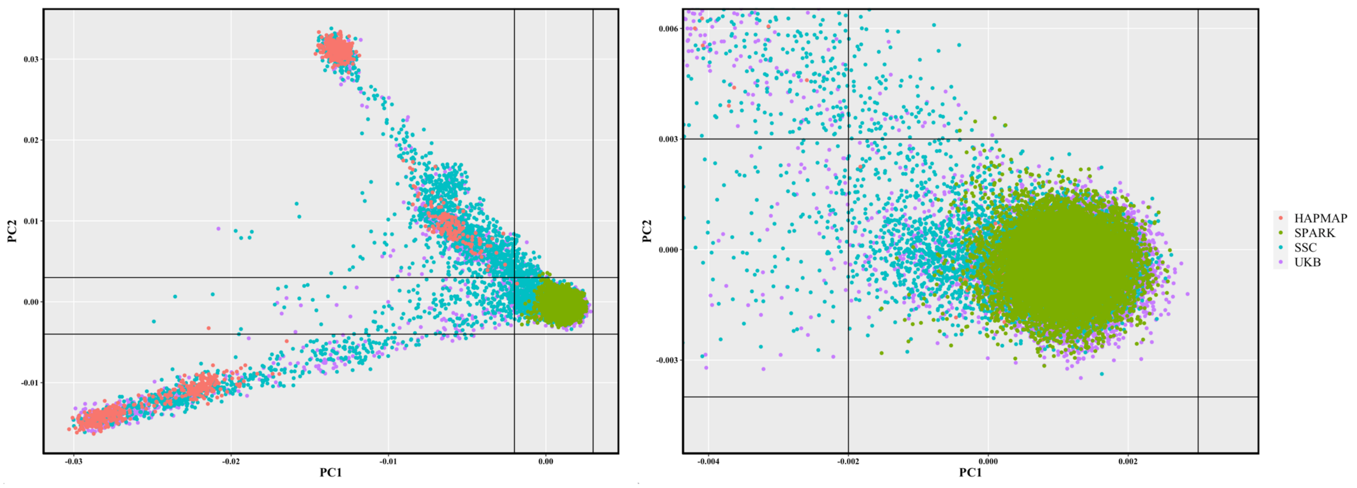


**Supplementary Figure 2. PCA of SPARK, SSC and UKB with HapMap**. See Online Methods for details

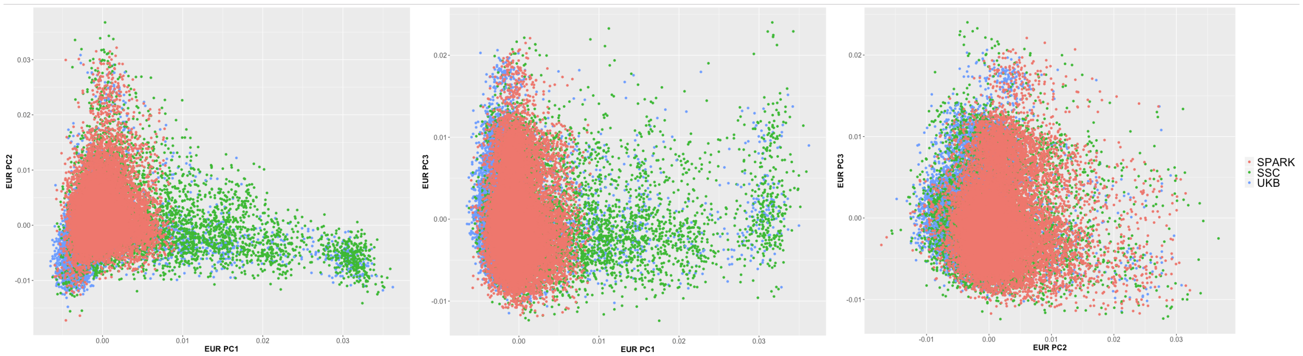


**Supplementary Figure 3. Within-European PCA of SPARK, SSC and UKB.** See Online Methods for details

### SPARK imputation methods

#### Ancestry Assignment

Self-reported demographic data were not available for the majority of SPARK participants, though existing data suggests that the racial and ethnic representation approximates that of the larger US population.[1,2] To determine which individuals were of European ancestry, we first restricted to a maximally unrelated ($\hat{\pi} < 0.09375$; midpoint between 3rd and 4th degree relatives) set of pedigree-reported founders as defined by PRIMUS ($n$ = 13,976).[3] We then performed[4] PCA via EIGENSOFT[5,6] on this sample after combining with those in the Human Genome Diversity Project (HGDP).[7,8] We used the HGDP sample in order to capture the full axes of ancestral variation within the SPARK sample. For the purposes of PCA, only variants passing a strict set of Ricopili QC measures (missingness < 5%, HWE $P > 1.0 \times 10^{-3}$, strand-unambiguous, and not in regions of high LD such as the MHC and chr8 inversion) were used, pruned to be pairwise independent at $r^2 < 0.2$. Additionally, 70 SNPs with allele frequency differences of > 0.2 between SPARK and HGDP self-reported EUR samples were removed. Non-founders were projected into the PC space of unrelated founders and the HGDP sample using hwe_normalized_pca in Hail (https://hail.is/). ADMIXTURE[9] was used in order to identify ancestral subpopulations within the joint SPARK + HGDP sample described above; cross-validation suggested the presence of 5 subpopulations. Individuals were labelled as having primarily EUR ancestry ($n$ = 17,098) if their ancestral makeup, as determined by ADMIXTURE, was 85% or greater from Population 0. Population 0 was determined to be the EUR subpopulation as it contained a high prevalence of HGDP EUR and self-reported SPARK White/Caucasian relative to other HGDP or other self-reported ancestry, respectively.

#### Pre-imputation QC

Upon restricting to individuals of primarily EUR ancestry, we undertook both sample and variant-level QC procedures consistent with the Ricopili and picolili standards. Samples were removed for the following reasons: missingness rate > 0.02 ($n$ = 71), absolute $F_{het}$ homozygosity rate > 0.2 ($n$ = 2), Mendelian error rate > 0.02 ($n$ = 0), sex check errors ($n$ = 14), and cryptic relatedness ($\hat{\pi}$ > 0.09375 across families; $n$ = 46). All self-reported pedigrees were confirmed via genetically derived kinship coefficients. Variants retained for inclusion were required to have missingness < 0.02; absolute differential missingness between cases and controls < 0.02; Mendelian error rates < 0.01; and HWE $P > 1.0 \times 10^{-10}$ in founder cases, HWE $P > 1.0 \times 10^{-6}$ in founder controls, and HWE $P > 1.0 \times 10^{-10}$ in all founders. Post-QC, 16,965 samples and 557,368 variants remained for imputation.

#### Imputation

Autosomes were imputed to the Haplotype Reference Consortium (HRC)[10] reference panel using SHAPEIT[11] and IMPUTE2[11,12] in the picopili pipeline (https://github.com/Nealelab/picopili). Phasing was performed using SHAPEIT including its duoHMM algorithm, which uses pedigree information when available for more accurate results.[13] Best-guess genotypes were called for autosomal SNPs (minimum posterior probability > 0.8) and subsequently filtered to SNPs with missingness < 0.02, INFO > 0.6, and MAF > 0.005, for a final total of 7,124,628 SNPs with a genotyping rate of 0.995 across 16,965 samples.
